# Introducing *SoNHR* – Reporting guidelines for social networks in health research

**DOI:** 10.1101/2023.04.19.23288797

**Authors:** Douglas A. Luke, Edward Tsai, Bobbi J. Carothers, Sara Malone, Beth Prusaczyk, Todd B. Combs, Mia T. Vogel, Jennifer Watling Neal, Zachary P. Neal

## Abstract

**Objective:** The overall goal of this work is to produce a set of recommendations (*SoNHR* – Social Networks in Health Research) that will improve the reporting and dissemination of social network concepts, methods, data, and analytic results within health sciences research.

**Methods:** This study used a modified-Delphi approach for recommendation development consistent with best practices suggested by the EQUATOR health sciences reporting guidelines network. An initial set of 28 reporting recommendations was developed by the author team. A group of 67 (of 147 surveyed) experienced network and health scientists participated in an online feedback survey. They rated the clarity and importance of the individual recommendations, and provided qualitative feedback on the coverage, usability, and dissemination opportunities of the full set of recommendations. After examining the feedback, a final set of 18 recommendations was produced.

**Results:** The final SoNHR reporting guidelines are comprised of 18 recommendations organized within five domains: *conceptualization* (how study research questions are linked to network conceptions or theories), *operationalization* (how network science portions of the study are defined and operationalized), *data collection & management* (how network data are collected and managed), *analyses & results* (how network results are analyzed, visualized, and reported), and *ethics & equity* (how network-specific human subjects, equity, and social justice concerns are reported). We also present a set of exemplar published network studies which can be helpful for seeing how to apply the SoNHR recommendations in research papers. Finally, we discuss how different audiences can use these reporting guidelines.

**Conclusions:** These are the first set of formal reporting recommendations of network methods in the health sciences. Consistent with EQUATOR goals, these network reporting recommendations may in time improve the quality, consistency, and replicability of network science across a wide variety of important health research areas.

## Introduction

Despite the dominance of the medical model of health in the 20th and 21st centuries[1], we have always understood that health is very much socially determined. Family and peer influences on smoking and diet; the role of social support on longevity and quality of life; social class and income inequality influencing access to health care; social isolation as a risk factor for depression and suicide; relational and structural factors shaping the course of pandemics – these are just some of the examples of how social factors are involved with individual, community, and population health [2].

Network science is the use of relational and structural theories to study network representations of complex social and physical systems. Although the roots of network science go back over a hundred years, its application within the health sciences is more recent. Driven by theoretical advances [3], modern computational power, and the increased availability of socially structured health data [4], the application of network designs and analytic methods within the health sciences has increased dramatically in the past few decades [5]. For example, Figure 1 shows the increase in network analytic studies over the past 20 years—with health-related network studies currently accounting for as much as 26% of all network publications.

**Fig 1.**
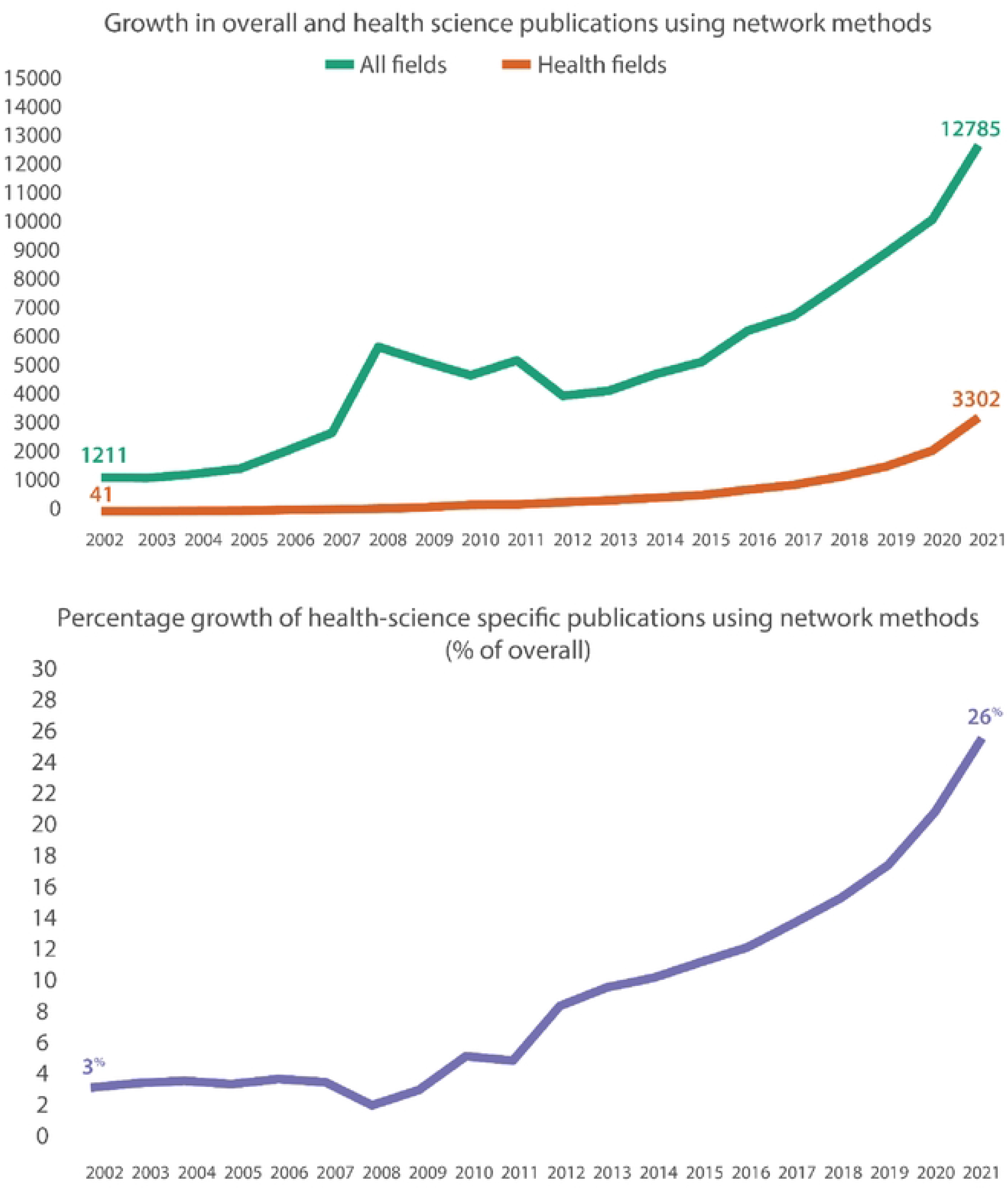
While network analysis studies have grown in general over the past two decades, the percentage of those specifically from health fields has grown from just 3% in 2002 to 26% in 2021. (The data for this figure come from SCOPUS searches for ‘network analysis’ first overall and then limited to publications focusing on health fields.)

Modern health science has benefited greatly from the reporting guidelines movement, where methodological, analytical, and reporting best practices are used to improve research quality, validity, and impact [6,7]. Numerous reporting guidelines exist across the social and health sciences that help guide practice and reporting of clinical trials (i.e., CONSORT), systematic reviews (PRISMA), epidemiologic studies (STROBE), and implementation science studies (StaRI), to name just a few. However, until now there has not been a similar set of guidelines for reporting the results of network studies in the health sciences. This is an important gap to fill, most directly because of the prevalence of network studies, as suggested above [8]. However, a set of network reporting best practices will also benefit a notable training gap among network analysis professionals. Training in network science and analytic methods is still not regularly featured in most medical schools, schools of public health, and even many social science departments.

The overall goal of this work is to produce a set of recommendations that will improve the reporting and dissemination of social network concepts, methods, data, and analytic results within health research. These recommendations, called Social Networks in Health Research (SoNHR), focus specifically on *reporting*, not on how to do better network science. There already exist many excellent network analysis texts, for example, those by Scott (2017) [9] or Borgatti, Everett, and Johnson (2018) [10]. And although the primary emphasis is on reporting network studies in peer-reviewed scientific publications, we anticipate that these recommendations will be useful in other contexts including evaluation reporting, and in training and teaching. The paper is structured in three parts: first, we describe the process we took for developing the network reporting guidelines. Second, we present and discuss the recommendations themselves. We then conclude with some recommendations on how to use the recommendations in different contexts and present some helpful resources including examples of the recommendations in practice.

## Methods

### Overview

This is a guidelines development study with the main goal of producing a set of reporting recommendations for studies using network data and methods in the health sciences. The general methodological approach we used for developing these recommendations was a modified expert panel consensus process, as recommended by EQUATOR (Enhancing the QUAlity and Transparency Of health Research; https://www.equator-network.org/) and implemented by other reporting guidelines development teams (e.g., Pinnock, et al., 2017) [11]. This study was approved by the Institutional Review Board of Washington University in August 2021 (ID: 202108053).

### Development of initial set of recommendations and expert panel survey

An initial set of network reporting recommendations was developed by the author team, based on an informal review of the network science and health science literatures, as well as their experience using network methods. Collectively, the author team has decades of experience conducting network studies, disseminating network studies to health and social science audiences, teaching and training students in network methods, publishing network methods texts, and reviewing grants and manuscripts using network and systems science methods. In this development phase, we produced a set of 28 candidate recommendations, grouped into five categories: 1) conceptualization, 2) operationalization, 3) data collection & management, 4) analyses & results, and 5) ethics & equity. These categories broadly represent the phases of empirical research, along with an overarching category focusing on ethical and equity aspects of network science [12].

Originally, we planned to convene network science experts to provide input and feedback on the recommendations. However, because of COVID-19 we switched to an online survey approach. The survey was developed in Qualtrics and focused on two types of feedback. Usability ratings were assessed using two items: *importance* of the recommendation, and *clarity* of the recommendation. Importance was defined as “the degree to which you feel that the particular guideline is critical for researchers to understand and follow in their empirical studies which apply SNA methods” and was rated on a 1 (Not at all Important) to 5 (Very Important) scale. Clarity was defined as “the degree to which the wording of the guideline clearly communicates to researchers using these reporting guidelines the type of information needed to satisfy the guideline” and was rated on a 1 (Not at all Clear) to 5 (Very Clear) Likert scale.

In addition to these quantitative items, participants were asked to respond to a series of open-ended questions focusing on general improvements to the guidelines, suggestions for disseminating the guidelines, and how they might use the guidelines in their own work. Appendix A contains the expert panel feedback survey, including the initial set of 28 recommendations.

### Participants

Our goal was to identify individuals who were experts in social network analysis applied to health contexts. To that end, we identified five stakeholder groups that should be represented: experts in applied health research, social network methodologists, network methods instructors, journal editors, and funders. Using these groups as guiding principles, we identified individuals who authored well-known papers, books, and analysis software; colleagues from federal funding agencies (NIH, CDC, etc.); participants in network analysis trainings (e.g., Systems Science for Social Impact); and members of the International Network for Social Network Analysis (INSNA). We invited a total of 147 people to participate, of whom 67 responded for a response rate of 45.6 %. The survey ran from December 2021 through April 2022. Responses were treated as confidential; specific identifying information was stripped from the data files prior to analyses.

Participant characteristics are presented in Tables 1 and 2. The participants as a group were experienced network and health scientists, working across a wide variety of fields and research areas. Over 80% of the participants had at least six years of experience using network designs and methods, with over 20% having at least two decades of experience. Nearly all participants had published network papers and reviewed network-focused papers or grants during the past five years (94% and 88%, respectively). The participant group included ten members who have worked in an organization that funds network research activities.

**Table 1.** Professional characteristics of expert panel members. (Percentages can add up to more than 100% because of multiple choice options.)

|  | Frequency | Percentage |
| --- | --- | --- |
| *Network Research Experience* |  |  |
| 20+ years | 14 | 21.5 |
| 11-20 years | 22 | 33.8 |
| 6-10 years | 18 | 27.7 |
| 1-5 years | 11 | 16.9 |
| *Network Science Activities (past 5 years)* |  |  |
| Published paper | 61 | 93.8 |
| SNA professional organization | 58 | 89.2 |
| Reviewed SNA papers/grants | 57 | 87.7 |
| Taught SNA | 48 | 73.8 |
| Worked in org. funding SNA | 10 | 15.4 |
| *Primary Disciplinary Focus* |  |  |
| Public health | 38 | 58.5 |
| Sociology | 29 | 44.6 |
| Statistics, Math, CS | 13 | 20.0 |
| Organizational research | 13 | 20.0 |
| Psychology | 11 | 16.9 |
| Medicine | 8 | 12.3 |
| Social Work | 7 | 10.8 |
| All others (15) | 31 | 47.6 |
| *Health Science Applications* |  |  |
| Implementation science | 25 | 38.5 |
| Health disparities, equity, justice | 25 | 38.5 |
| Community-focused | 22 | 33.8 |
| Methods development | 21 | 32.3 |
| Program evaluation | 19 | 29.2 |
| Health services | 18 | 27.7 |
| Mental health | 17 | 26.2 |
| Epidemiology | 16 | 24.6 |
| Health promotion | 15 | 23.1 |
| Addiction, substance use | 14 | 21.5 |
| Health policy | 13 | 20.0 |
| Children, youth, families | 13 | 20.0 |
| Older adults | 12 | 18.5 |
| Cancer | 11 | 16.9 |
| Infectious disease | 11 | 16.9 |
| All others (17) | 67 | 101.5 |

**Table 2.**
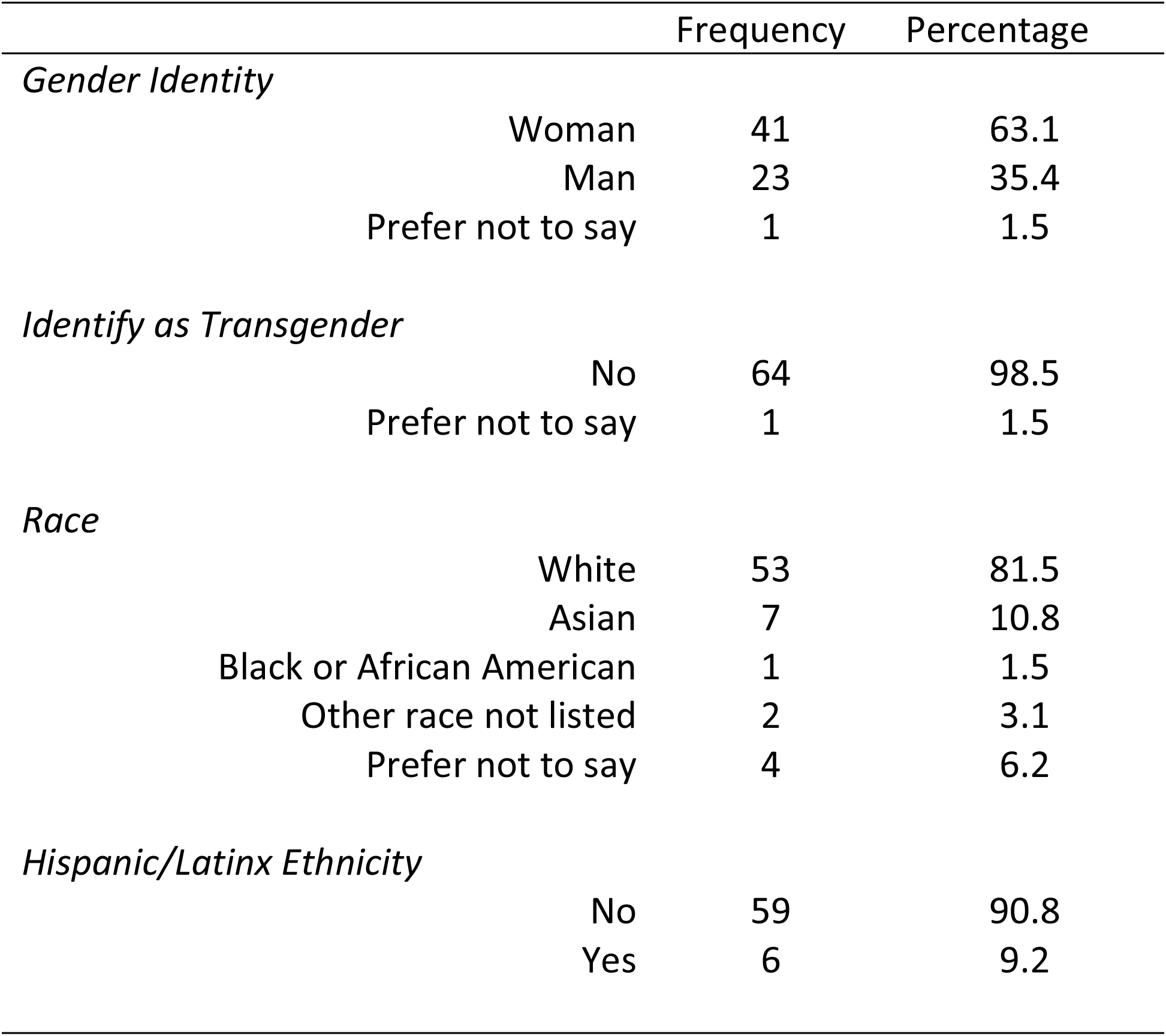
Demographic characteristics of expert panel members.

|  | Frequency | Percentage |
| --- | --- | --- |
| *Gender Identity* |  |  |
| Woman | 41 | 63.1 |
| Man | 23 | 35.4 |
| Prefer not to say | 1 | 1.5 |
| *Identify as Transgender* |  |  |
| No | 64 | 98.5 |
| Prefer not to say | 1 | 1.5 |
| *Race* |  |  |
| White | 53 | 81.5 |
| Asian | 7 | 10.8 |
| Black or African American | 1 | 1.5 |
| Other race not listed | 2 | 3.1 |
| Prefer not to say | 4 | 6.2 |
| *Hispanic/Latinx Ethnicity* |  |  |
| No | 59 | 90.8 |
| Yes | 6 | 9.2 |

### Development of final set of network reporting recommendations

Data from the expert panel survey were summarized and analyzed. Specifically, descriptive statistics and distributional characteristics of the importance and clarity items were examined to identify recommendations that could be dropped or recommendations that needed improvement. Thematic summaries were prepared for the open-ended questions and were also used to help differentiate between strong and weaker recommendations.

## Results

### Recommendations development

The quantitative results of the preliminary network reporting recommendations from the Delphi survey are presented in Table 3. In general, the preliminary recommendations received high scores on *importance* and *clarity.* The operationalization recommendations scored the highest, with average importance ratings of 4.7 (out of 5) and clarity ratings of 4.6. The five preliminary visualization recommendations scored lowest in importance (mean = 3.5). Closer examination of the visualization ratings suggested that participants fell into one of two categories—some felt these recommendations were quite important, but a substantial number clearly felt that visualizations in general were less important in network science. The detailed, item-level survey responses (including dot-plots) are available in Appendices B and C.

**Table 3.** Summary of importance and clarity ratings of preliminary set of network reporting recommendations.

| Domain | No. of Recs. | Importance |  | Clarity |  |
| --- | --- | --- | --- | --- | --- |
|  |  | Mean | Range | Mean | Range |
| Conceptualization | 3 | 4.3 | 4.1 – 4.6 | 4.2 | 3.7 – 4.4 |
| Operationalization | 4 | 4.7 | 4.4 – 4.9 | 4.6 | 4.4 – 4.7 |
| Data Collection & Management | 6 | 4.3 | 4.0 – 4.6 | 4.1 | 3.8 – 4.5 |
| Analysis & Results - Description | 2 | 4.3 | 4.2 – 4.4 | 4.1 | 4.0 – 4.2 |
| Analysis & Results - Visualization | 5 | 3.5 | 3.1 – 3.8 | 4.1 | 3.9 – 4.3 |
| Analysis & Results – Modeling & Simulation | 4 | 4.4 | 4.2 – 4.7 | 4.2 | 3.9 – 4.4 |
| Ethics & Equity | 4 | 4.2 | 3.8 – 4.6 | 4.3 | 3.8 – 4.5 |

The responses to the open-ended questions indicated general enthusiasm for the network reporting recommendations (e.g., “These are awesome and well needed. Bravo!”), but more importantly included many suggestions on how to improve individual recommendations as well as their overall structure. Participants also provided numerous suggestions on how to disseminate the guidelines as well as how they might use the recommendations in their own work (see Discussion below).

Taking the importance, clarity, and qualitative feedback into account, the team revised the recommendations by dropping, rewriting, and reordering items. Items were dropped if they received lower importance or clarity ratings, or if on further reflection they seemed to be redundant with other items or were too narrow in scope. For example, one of the preliminary 28 guideline recommendations that was dropped was, “For valued networks, describe reconciliation of conflicting values when provided by both members of the dyad.” This item was overly complicated (receiving the lowest clarity score in its group) and it applies to relatively few types of network studies. The main structural change we made was to combine two separate recommendation sections (*Network Descriptio*n and *Network Visualization*) into one section.

### Final reporting guidelines for social networks in health research (SoNHR)

Table 4 presents our final set of 18 recommendations for reporting social network methods and results, particularly within health sciences research (SoNHR). These recommendations are not a formal checklist; we do not expect all these recommendations to apply to every specific network paper. However, the use of these recommendations will help ensure that network methods and results are reported as clearly as possible, benefiting future research.

**Table 4.**
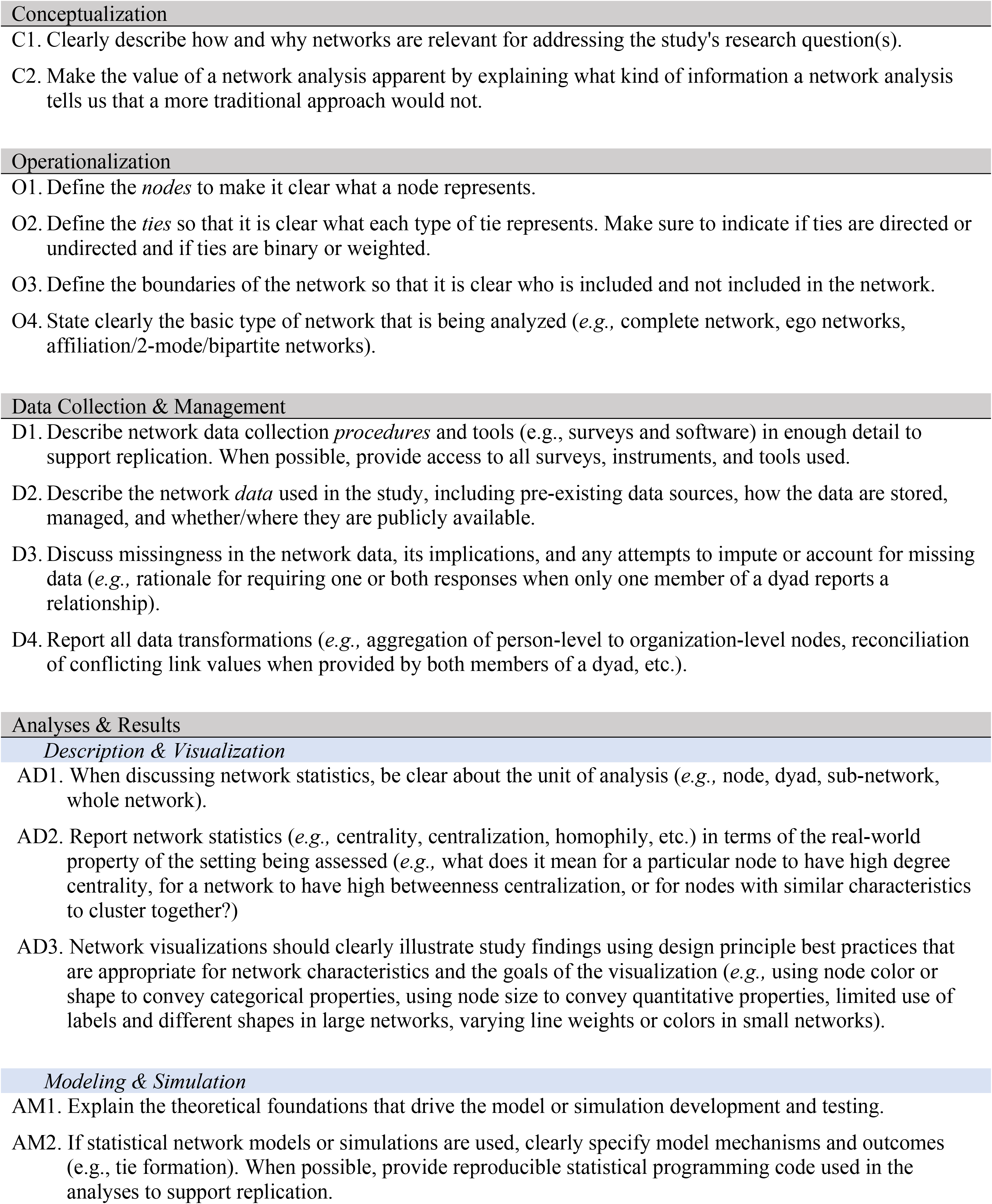
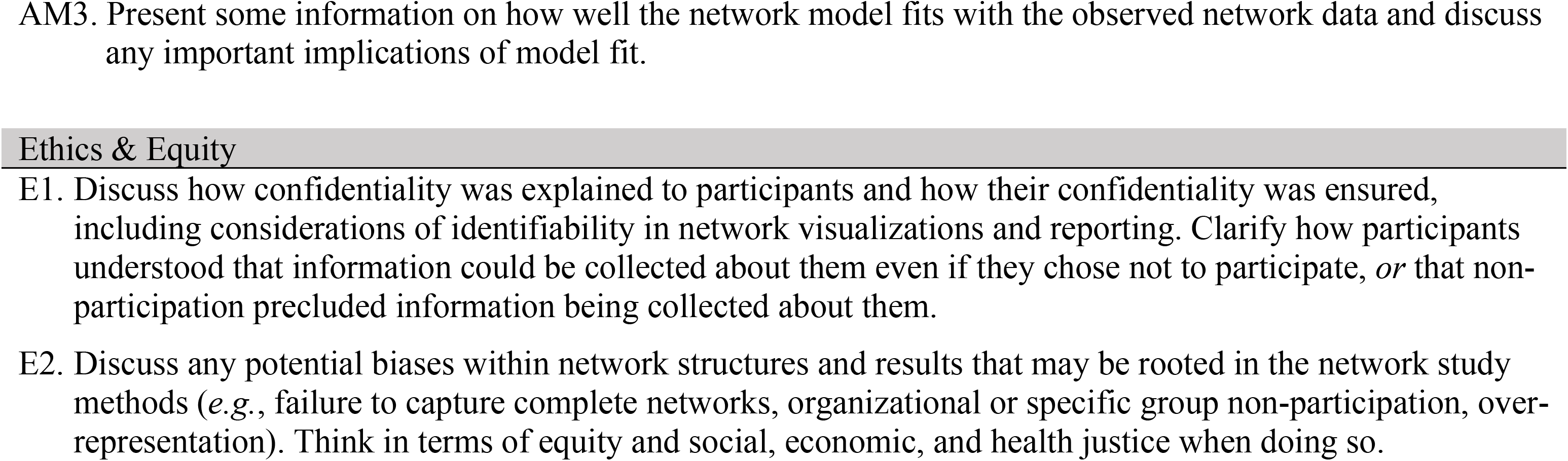
Final reporting guidelines - Social Networks in Health Research (SoNHR).

### Conceptualization

The first two recommendations refer to the basic *conceptualization* of the research project as reported in the publication. Network studies are inherently structural or relational; they concern themselves with how social objects (e.g., persons, organizations) relate to one another, or how social structures shape the flow of physical or social things such as viruses, information, money, behavior, etc. [13]. The first recommendation is to make the structural and relational conceptualizations that drive the research questions clear to the reader. This provides the basic rationale for using network methods in the first place. The second recommendation is similar but focuses on the network methods themselves. How are these network methods used in the study necessary or advantageous, given the research questions driving the study?

### Operationalization

The second set of recommendations is focused on *operational definitions* of the various pieces of the network study, *i.e.,* who is in the network, what the ties represent, and what kind of network is being analyzed. Definitions of the nodes are usually straightforward, but they may need additional details if the nodes are not persons (e.g., organizations), or in the case of 2-mode networks, where both types of nodes need to be clearly defined. For example, in an infectious disease contact tracing study, one type of node would be people who are infected, and the other type of node would be defined as a specific physical location where people come into contact with one another [14].

Defining the network ties used in a study often requires careful attention to detail. At a minimum, the description of network ties should include a statement of what the tie represents (e.g., contact frequency, trust, sexual contact) and how it was measured (e.g., self-report using a 5-point scale with each anchor point defined). This is particularly important when more than one type of tie is collected and analyzed in the study. Moreover, once ties have been defined, it is a good practice to refer to that type of tie specifically (e.g., ‘friendship tie’) rather than a more generic term like ‘ties,’ or ‘connections.’ Otherwise, this can lead to the impression by the reader that a network has only one type of relationship among network members when any network contains many different kinds of social relationships, whether measured in the study or not.

It is always important to describe who is in the network, and, conversely, who is not in the network. Thus, the boundaries of the network should be clear, especially for complete network studies [15]. And this leads to the last recommendation in this section, which is a clear description of the type of network being analyzed. First, state whether this network is *complete*, where most or all members of a boundary-defined network are included (e.g., ‘all cancer healthcare providers in St. Louis County); or *ego-centric*, where networks are constructed from the perspective of individual members or ‘egos’ (e.g., personal support networks of people in a study of the effects of social support). In addition, it is often important to define the network in terms of the types of ties—are the ties directed or non-directed, and are the ties valued or binary?

### Data Collection & Management

The third set of recommendations concerns reporting of *data collection and management* details. Network data are quite different from the data used in the vast majority of other health and social science research. They are fundamentally relational, which implies different kinds of data collection, management, and subsequent analytic practices. Therefore, authors may need to provide more information on these data management steps than they normally would. Taken together, these data management recommendations support future replication. They also suggest network-specific data issues. For example, missing data in network studies represent more serious threats to accurate interpretation and bias [16,17], so it is important to describe how missing data are handled in any network study. Furthermore, a common data issue in survey-based network analysis is when individuals in a dyad give conflicting information about a shared tie. If this is the case, then authors should report how that conflicting information is resolved [18]. In addition, enough details on any network data transformation (e.g., turning 2-mode into 1-mode network data [14]) should be presented to support full understanding and potential future replication.

### Analyses & Results

The first set of recommendations under analyses and results helps ensure that readers will fully understand the results coming out of basic *descriptive network analysis* studies. First, make sure to clarify what the network *unit-of-analysis* is for any particular set of network statistics. Readers may not fully appreciate how network results can focus on individual network members, pairs of network members, subnetworks, or complete network characteristics. Second, it is very important to convey how a particular network statistic measures or captures an underlying behavioral, structural, or relational characteristic of the network or network member. For example, in a communication network study, do not simply report the technical formula for a statistic such as network *centralization,* but describe how this measure assesses the extent to which the communication structure is more or less hierarchical in nature. Finally, many network studies employ visualizations, so it is important to design them in such a way that it is clear how the visualization supports or reveals the underlying network characteristic or analytic result. This is likely to require both general information visualization skills (e.g., Tufte, 2001), as well as network-specific graphic design principles (e.g., Shneiderman & Dunne, 2012; Ognyanova, 2021).

The second section of recommendations under analyses and results refers to good practices for reporting results from *network modeling or simulation* studies. These types of studies move beyond simple network descriptions and visualizations to using data and theory to pose and test hypotheses about network structures and influences. Taken together, these three recommendations will help ensure that readers will understand the theory or framework that drives the network modeling analyses, will have enough technical information to support further research replication using these models, and appreciate how well the network model or simulation fits with the observed network data [22].

### Ethics & Equity

The final set of recommendations refers to special considerations around *ethics and equity* in network studies. Standard non-network surveys and similar methods of data collection allow for anonymity and confidentiality. However, when collecting data on networks anonymity is not possible, as the individuals or organizations in the network must be named to create ties in subsequent analyses. Even if a person or organization chooses not to participate in a network study, they might be named by others during data collection. Offering informed consent or even *truly informed consent* for participants in network studies is a promising practice to ensure confidentiality [12,23]. As part of this process, those choosing not to participate should be aware that they may be named by others. Omitting non-consenters in a network study is dangerous and can greatly impact network structures, analyses, results, and interpretations [24]. More appropriate strategies for maintaining confidentiality are to anonymize data as soon as possible after data collection and to reduce the number of people who have access to the data [23,25].

Equity is an essential consideration in all scientific research, and in network analysis its importance is amplified. As stated above, maximizing response rates and capturing complete networks are also especially important in network studies. It is important to identify all who influence or are affected by the substantive area of the network under consideration, including historically underrepresented types of network members such as patients in individual networks or relatively small agencies in organizational networks. Researchers should monitor recruitment and look for patterns in non-responses. For example, one study found that network studies of sex workers commonly underrepresented men in their roles as sex workers and designed a purposeful sampling strategy more inclusive of them [26]. Similar to the ethical concerns around reporting incomplete network study findings, the underrepresentation of certain groups or organizations can distort results, or even worse, exacerbate existing inequities through exclusion.

## Discussion

The SoNHR (Social Networks in Health Research) guidelines were developed to promote the clear and comprehensive reporting of network studies in the literature among health researchers. Already a well-established theoretical and methodological frame, the use of network science in health research continues to increase. However, without reporting guidelines, the utility, replicability, and impact of network science studies are diminished. These guidelines were created through an iterative, expert consensus building process and the resulting recommendations can be applied broadly across health research studies, including those in the clinical, social science, community, and population/public health research fields.

### Suggested audiences and supporting resources

These guidelines have the potential to benefit at least five distinct audiences – instructors, authors, journal editors, reviewers, and readers – that capture the full process of scientific knowledge generation and dissemination.

First, these guidelines offer *authors* recommendations about how to report the design and results of their health-related social network research clearly and consistently. We anticipate this is especially helpful and important because social network methods are both diverse and rapidly developing, which can lead to confusion among authors about what essential aspects to report. Although the guidelines themselves offer specific recommendations, Table 5 provides examples of published research that have followed these recommendations, and thus that can serve as exemplars for authors. Although not exhaustive, these examples represent a collection of “best practices” for each of the recommendations and illustrate how each of the recommendations can be put into action.

**Table 5.**
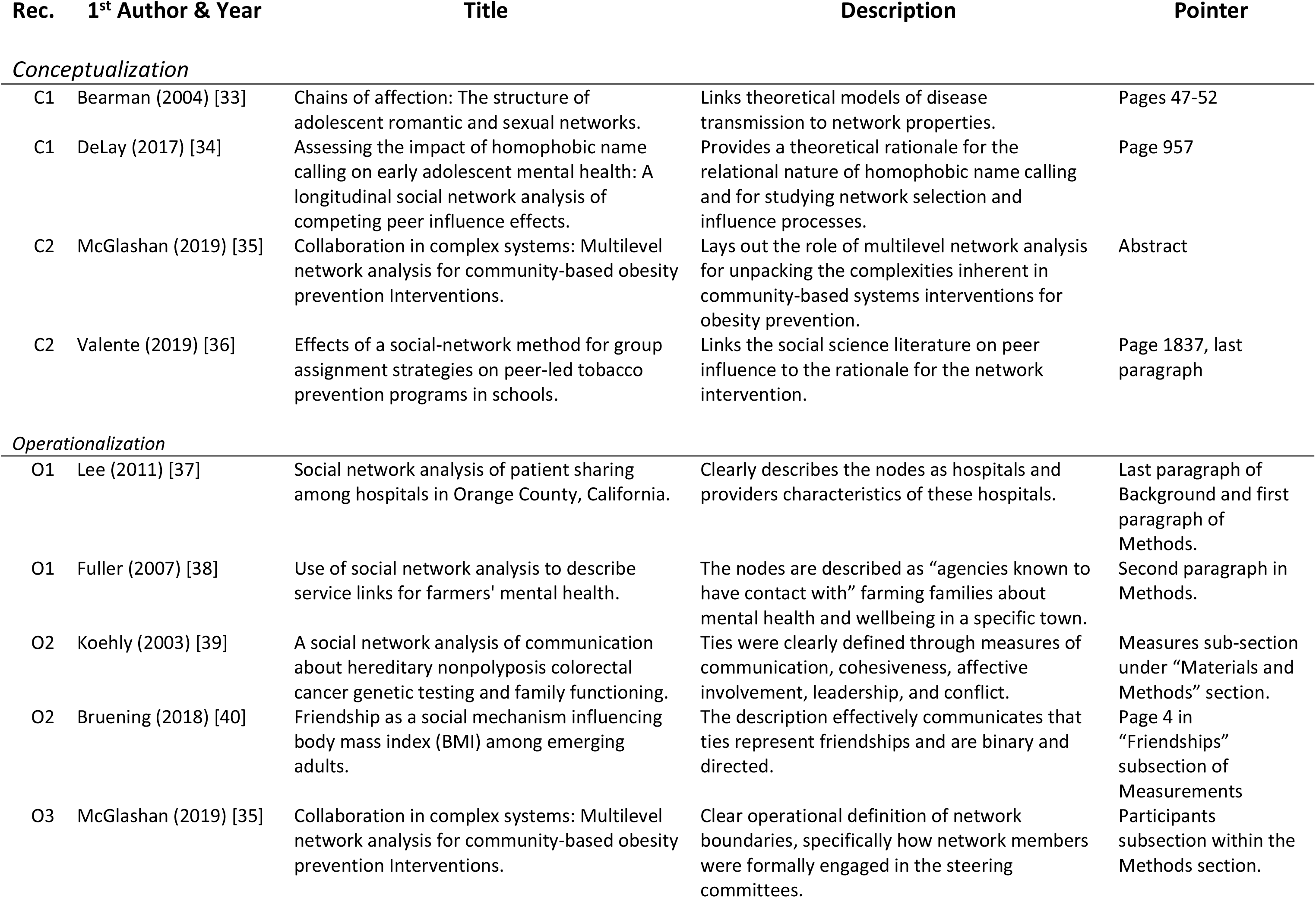
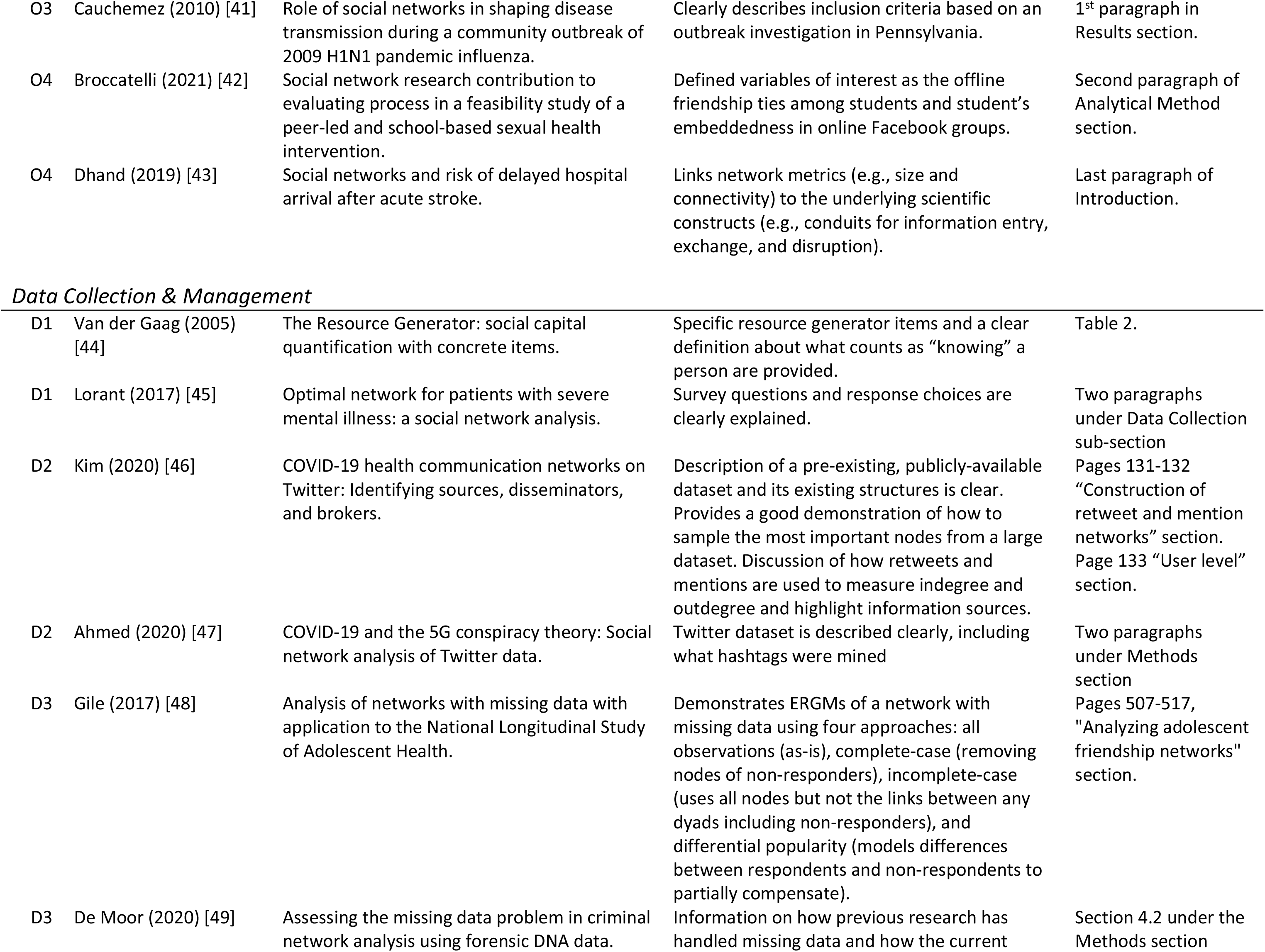
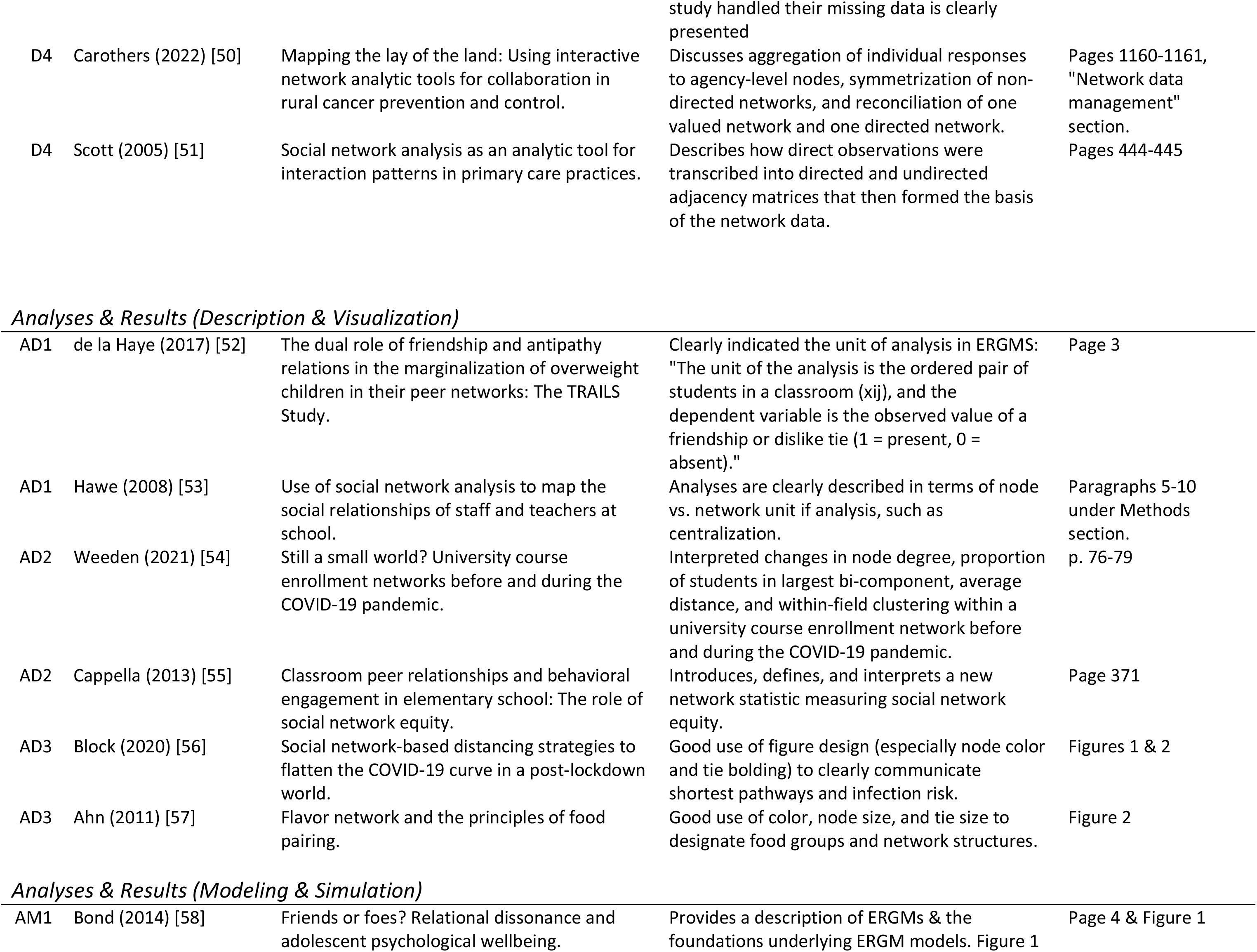
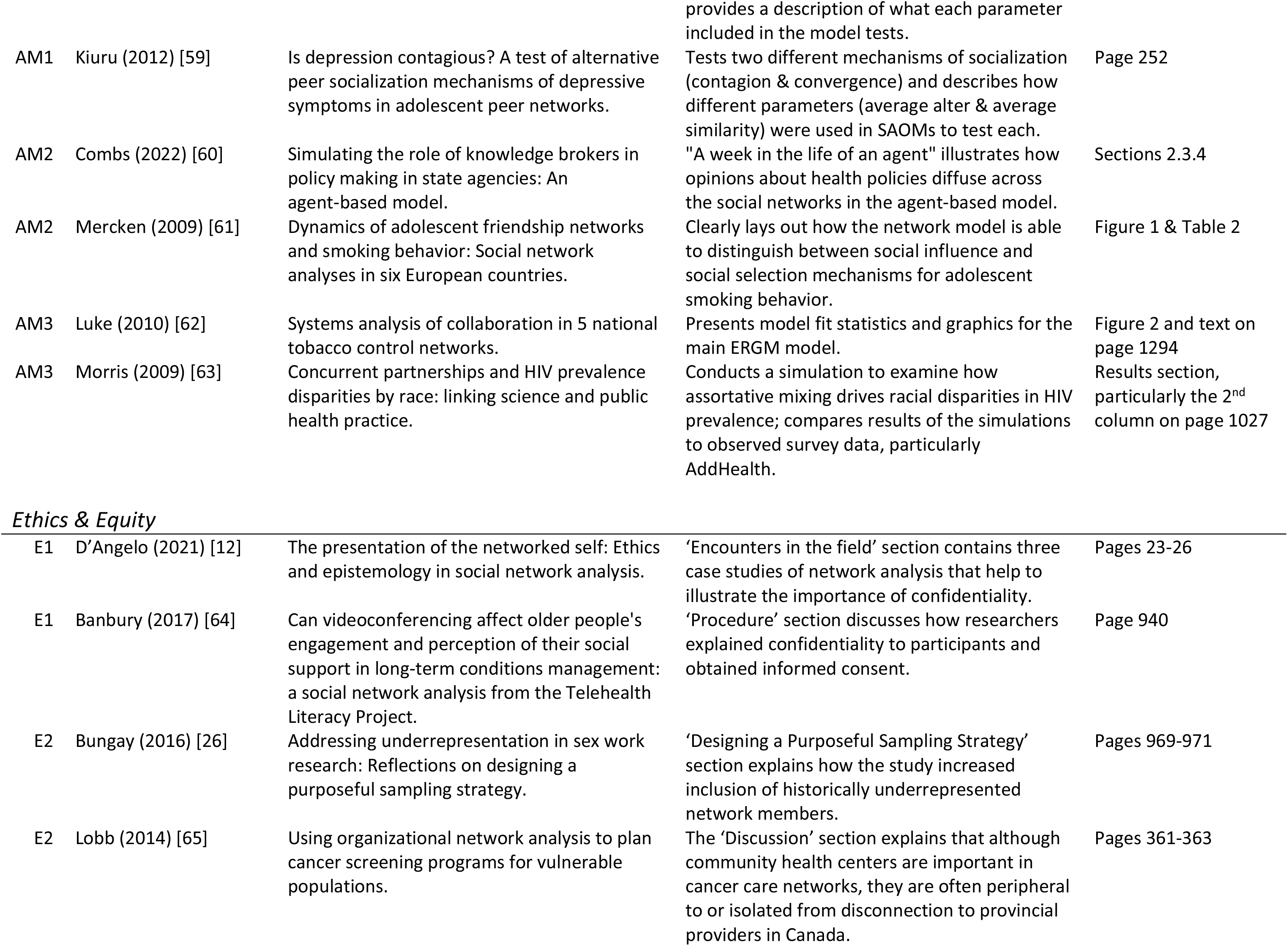
Non-exhaustive list of published network studies demonstrating network reporting recommendations.

Second, these guidelines can be used by *instructors* of social network coursework to inform the topics covered in such courses, and by their students to rapidly develop an understanding of the most important features of social network research. For example, modules in an introductory course on social network analysis could roughly follow the major headings of the guidelines, while individual recommendations under each of these headings could serve as learning objectives within these modules.

Third, these guidelines can serve as a helpful checklist for *reviewers* when evaluating health-related social network research. Reviewers can use the guidelines and individual recommendations to ensure the completeness of their evaluations of social network studies. In addition, following these guidelines to evaluate social network research may improve the consistency between reviewers, which is often quite low [27], as they provide a common set of reporting standards to evaluate.

Fourth, these guidelines can be required as reporting standards by *journal editors* who publish health-related social network research to ensure the standardized reporting of social network methods and results. This type of editorial requirement is not without precedent. For example, many journal editors already require reporting guidelines developed for other methods including PRISMA for systematic reviews and meta-analyses [28], JARS-QUAL for qualitative research [29], JARS-QUANT for quantitative research [30], and CONSORT for randomized trials [31].

Finally, these guidelines have benefits for *readers* of health-related social network research. When implemented, these guidelines should improve the detail, clarity, and transparency of social network methods and results for readers. Additionally, when put into practice, these guidelines should ensure that critical methodological and analytic details are provided in social network papers, making it easier to re-use published social network research in meta-analyses and systematic reviews.

### Strengths, gaps, and next steps

This is among the first set of reporting guidelines for social network science concepts, methods, and results. Although a number of network *data* formatting and reporting guidelines exist (e.g., Bagrow, 2022) [32], to our knowledge there are no guidelines focusing on best reporting practices in scientific dissemination, especially within health research. These guidelines are being published in EQUATOR, which will increase their visibility and longevity and hopefully enhance their effectiveness. Finally, these reporting guidelines include equity and ethical considerations, which are important, require special consideration in network science, and have previously been underdiscussed.

Although we developed these reporting guidelines to be broadly applicable to many kinds of network studies, they may be less helpful (or need to be adapted) for some kinds of network study designs or analytic frameworks (e.g., purely qualitative network studies, ego-centric designs, dynamic analyses, etc.). Similarly, the scope of these recommendations is somewhat high-level, we deliberately left out extremely specific reporting suggestions (e.g., guidance around the use of weighted ties in network visualizations) that would not be applicable to many network studies. Finally, for the expert panel consensus process, we may have missed some important viewpoints and distinct experiences that could have helped inform the network reporting guidelines. However, the relatively large, experienced, and diverse set of participants we ended up with (see Tables 1 & 2) somewhat mitigates that concern.

We will continue to work on disseminating the SoNHR guidelines beyond EQUATOR and then hosting these guidelines on relevant websites (including at <u>cphss.wustl.edu</u>). We also encourage interested partners to further spread the guidelines and share how they have used them (see above). In particular, case studies of the application of the network reporting recommendations would be useful for the field of network health research.

As we gain a better understanding of how social connections, structures, and dynamics are implicated in human disease processes, effective healthcare delivery, and promotion of public health, there will be a concomitant need for appropriate scientific frameworks, study designs, and analytic approaches. Network science and social network analysis are particularly well-suited for these types of health research. The network reporting guidelines presented here will be helpful for ensuring that the knowledge generated in these studies can have the broadest impact, both scientifically and socially.

## Data Availability

All relevant data are within the manuscript and its Supporting Information files.

## Acknowledgements

We are grateful for the contributions and expertise of our recommendations development expert panel. They were crucial for helping us develop and finalize the SoNHR recommendations.

## Supporting information

**S1 File. Appendix A.**

**S2 Table. Appendix B.**

**S3 Fig. Appendix C.**

**S4 File. Feedback Data**

**S5 File. Feedback Data Variable Dictionary**

## Notes

### Competing Interest Statement

The authors have declared no competing interest.

### Funding Statement

Research reported in this publication was supported by the following sources: - Implementation Science Centers in Cancer Control (National Cancer Institute and the Barnes Jewish Hospital Foundation P50CA244431 DAL, ET, SM, BP) - Washington University Institute of Clinical and Translational Sciences (National Center for Advancing Translational Sciences UL1TR002345 DAL, BJC, TBC) - Centers for Diabetes and Translational Research (National Institute of Diabetes and Digestive and Kidney Diseases, P30DK092950 DAL) - Improving Alzheimer’s Disease and Related Dementias Care in Rural Areas (National Institute of Aging K01AG071749 BP) - T32 Training Program (National Heart, Lung, and Blood Institute T32 HL130357 MTV) - T32 Training Program (National Cancer Institute T32CA190194 ET) The funders had no role in study design, data collection and analysis, decision to publish, or preparation of the manuscript.

### Author Declarations

The Human Research Protection office at Washington University in St. Louis deemed this project exempt (#202108053), and we obtained electronic informed consent from all participants.

